# Characterizing the Emergence of Medetomidine in Southern California Fentanyl Markets

**DOI:** 10.64898/2026.10.13.26365459

**Authors:** Adam J. Koncsol, Joseph R. Friedman, Caitlin A. Molina, Megan Grabill, Morgan E. Godvin, Ruby Romero, Elizabeth A. Samuels, Edward Sisco, Chelsea L. Shover

## Abstract

**Background:** Medetomidine, a potent sedative, is increasingly present in fentanyl-related hospitalizations and deaths. Multiple states have passed legislation to control xylazine—a pharmacologically similar sedative in the illicit opioid supply. Medetomidine may function as a highly potent substitute for xylazine with significant overdose risk and withdrawal potential. Medetomidine has previously been described mostly in eastern U.S. fentanyl markets. Here, we characterize medetomidine-containing drug samples obtained in Southern California 2023–2026.

**Methods:** Anonymous participants in Southern California voluntarily provided drug products to a community-based drug checking program. All samples underwent direct-analysis-in-real-time mass spectrometry (DART-MS). A subset was quantified by liquid-chromatography tandem mass spectrometry (LC-MS/MS). A smaller subset underwent enantiomeric analysis. We calculated other compounds co-present with medetomidine, the prevalence of medetomidine in fentanyl over time, the concentration of medetomidine, and the enantiomer makeup of medetomidine.

**Results:** The quarterly prevalence of medetomidine co-present with fentanyl rose from ≈6 % in May 2025 (month of first detection) to ≈25 % by August 2026. The average medetomidine concentration was 0.4 % mass fraction. The most common co-detected substances with medetomidine were fentanyl and adulterants such as lidocaine and xylazine. All medetomidine samples that underwent enantiomeric analysis (n = 13) were identified as racemic.

**Conclusion:** Medetomidine represents a rapidly emerging adulterant in the Southern California fentanyl supply. All samples assessed for enantiomer status were racemic medetomidine, suggesting they were not diverted from human pharmaceutical or medical supplies. Given its high potency, a small amount (<1.5%) is likely still a physiologically significant quantity of the drug.

**Highlights:**

- Medetomidine is an emerging adulterant in the Southern California fentanyl supply.
- The share of fentanyl with medetomidine rose from 6% in May 2025 to 25% in Aug 2026.
- Nearly all medetomidine samples were detected in the presence of fentanyl.
- Medetomidine was found in two samples of DMT obtained via the dark web.
- The mean concentration of medetomidine was 0.4 % mass fraction among 22 cases.
- All tested medetomidine was racemic, not diverted dexmedetomidine.

## Background

Medetomidine is a potent sedative approved by the U.S. Food and Drug Administration for use in veterinary medicine. An enantiomer of medetomidine, *dexmedetomidine*, is also used in humans as a sedative in hospital intensive care units (de Andrade Horn et al., 2024). Various emerging reports suggest that medetomidine is increasingly being found in the illicit fentanyl supply in North America. Medetomidine is pharmacologically related to xylazine—another sedative drug approved for use in animals that has significantly impacted U.S. illicit fentanyl markets (Gupta et al., 2023; Montero et al., 2022)—through the same neurochemical activation on α2-adrenoreceptors (de Andrade Horn et al., 2024). In animal studies, medetomidine has been estimated to be potentially up to 100 times stronger than xylazine (de Andrade Horn et al., 2024). Medetomidine has been detected alongside or as an apparent substitute for xylazine in fentanyl drug product and paraphernalia (Palamar & Krotulski, 2024; Sisco & Appley, 2023) and is increasingly co-present in illicit fentanyl-related hospitalizations and deaths (Palamar & Krotulski, 2024; Sood, 2025). Along with complicating overdose and overdose reversal, medetomidine, like xylazine, has been associated with development of tolerance and severe withdrawal syndrome that may be higher severity than that observed with xylazine alone (CDC, 2026; London et al., 2026; Ostrowski et al., 2025; Sood, 2025).

In the years since xylazine emerged in the illicit drug supply (Friedman et al., 2022), multiple states have passed or considered legislation to control xylazine (Sugarman et al., 2024). Contemporaneously, medetomidine has emerged as an increasingly prevalent adulterant throughout the East coast, Midwest, and other regions where xylazine has historically been present in the fentanyl supply. Xylazine has been documented in Southern California at a comparatively lower prevalence and concentration in samples of fentanyl drug products than in East Coast locations where it was initially popularized (Friedman et al., 2025). However, little is known about the extent to which medetomidine is present in West Coast opioid markets. We describe the prevalence and concentration of medetomidine-involved drug product samples identified in Southern California from February 2023 until August 2026 through the use of community-based drug checking data. To our knowledge, this represents the first study of its kind to detail the arrival of medetomidine on the West Coast and to characterize its concentration and chirality.

## Methods

Anonymous participants in Southern California voluntarily provided samples (approximately 5 mg) of drug product to a community-based drug checking program at four sites between February 2023 and August 2026. All samples were sent to the National Institute of Standards and Technology Rapid Drug Analysis and Research program (NIST RaDAR) for testing. While these laboratory methods have been previously described in depth (Appley et al., 2023; Friedman et al., 2025; Shover et al., 2025; Sisco et al., 2017), briefly, NIST RaDAR evaluated all samples included in the study with direct-analysis-in-real-time mass spectrometry (DART-MS) and a subset of samples with liquid-chromatography tandem mass spectrometry (LC-MS/MS). DART-MS methods assess the presence or absence of substances by analyzing quantities as small as nanograms of suspected drug product samples against a spectral library of over 1,600 reference standards which include most significant psychoactive compounds as well as many other bulking or cutting agents, precursors, byproducts, and other adulterants and emerging novel substances. The limit of detection for DART-MS is approximately 0.05 % mass fraction, though that is dependent on starting mass and sample complexity.

LC-MS/MS methods are able to quantitate (as a mass fraction) a panel of pre-specified compounds of specific drugs, precursors, and adulterants of interest (see **Supplemental Table A** for a full list). Concentrations reported by NIST RaDAR are the freebase form of the drug and therefore do not account for the weight of the salt. The values reported by LC-MS/MS quantitation have high sensitivity, specificity, and the precision of quantitation estimates is typically considered to be within 20 % of the reported concentration; e.g., a reported concentration of 1 % mass fraction would have an uncertainty interval from 0.8 % mass fraction to 1.2 % mass fraction. The limit of quantitation for LC-MS/MS is approximately 0.1 % mass fraction. Compounds below the limit of quantitation were imputed at 0.1 % mass fraction for the purposes of computing statistics, although they may be present at lower amounts.

A smaller subset of selected medetomidine-positive samples also underwent enantiomeric analysis using LC-MS/MS with a chiral column which can provide information on the chirality of medetomidine. This type of analysis began in January 2026 and was typically limited to five medetomidine-positive samples per month. This analysis distinguishes between dexmedetomidine, the enantiomer of medetomidine approved by the FDA for use as a sedative in humans, and levomedetomidine, an enantiomer that is not approved for human use. This delineation can help indicate whether medetomidine present in illicit drug supplies may be dexmedetomidine, levomedetomidine, or an equal, racemic, mixture of the two. These findings help provide insight on whether the substance is diverted from the human pharmaceutical supply.

During the study period, 3479 samples were submitted to the lab and underwent DART-MS analysis. Of those, 1100 were confirmed to contain fentanyl by DART-MS. Medetomidine has primarily been found as an adulterant in illicit fentanyl and was first detected in our drug checking program in May 2025. The denominator for fentanyl-specific sub-analyses was therefore restricted to n = 426 fentanyl-containing samples tested from May 2025 – August 2026. We calculated (1) the detection of other compounds co-present with medetomidine, (2) the concentration of medetomidine, (3) prevalence of medetomidine in fentanyl over time, and (4) the enantiomer makeup of medetomidine. The UCLA Institutional Review Board reviewed and approved this project (protocol IRB-22-0760 and IRB-22-1273) and determined some components of this work constituted public health surveillance. Sample analysis at NIST was approved by the NIST Research Protections Office (MML-2021-0328).

## Results

There were n = 32 samples which were confirmed to have medetomidine present via DART-MS. Medetomidine was first identified in our sample in Southern California in May 2025. **Figure 1A** and **Table 1** show the co-prevalent compounds of the individual 32 samples detected with medetomidine. The most commonly co-detected substances with n = 32 medetomidine were fentanyl (≈94 %, n = 30), fentanyl precursor chemicals such as 4-ANPP (≈88 %, n = 28) and phenethyl 4-ANPP (≈25 %, n = 8), and adulterants like lidocaine (≈25 %, n = 8) and xylazine (≈16 %, n = 5).

**Table 1:** Count and proportion of compounds co-detected with medetomidine.

| **Class** | **Compound** | **Count (n)** | **Proportion (%)** |
| --- | --- | --- | --- |
| **α<sub>2</sub> sedatives** | Medetomidine | 32 | 100% |
|  | Xylazine | 5 | 15.6% |
| **Fentanyl, fentalogs, and fentanyl precursors** | Fentanyl | 30 | 93.8% |
|  | 4-ANPP | 28 | 87.5% |
|  | Phenethyl 4-ANPP | 8 | 25.0% |
|  | 1-phenethylpiperidin-4-ol | 4 | 12.5% |
|  | Ethyl 4-ANPP | 3 | 9.4% |
|  | Norfentanyl | 3 | 9.4% |
|  | Fluorofentanyl | 1 | 3.1% |
|  | N-phenethyl-N-phenylpropionamide | 1 | 3.1% |
| **Anesthetics** | Lidocaine | 8 | 25.0% |
|  | Cocaine | 2 | 6.2% |
|  | Benzocaine | 1 | 3.1% |
| **Other drugs** | Acetaminophen | 3 | 9.4% |
|  | Levamisole | 3 | 9.4% |
|  | Caffeine | 2 | 6.2% |
|  | DMT | 2 | 6.2% |
|  | MIPT | 2 | 6.2% |
|  | 5-MeO-DMT | 2 | 6.2% |
|  | Methamphetamine | 1 | 3.1% |
| **Others** | Mannitol | 8 | 25.0% |
|  | DEET | 1 | 3.1% |
| **Total** |  | **32** | **100%** |

Note: N = 32 medetomidine-involved samples and co-detected compounds confirmed present on DART-MS. Mannitol may be underrepresented here as DART-MS methods generally have difficulty detecting sugars. Data collected February 2023 to August 2026 in Southern California.

**Fig. 1A:**
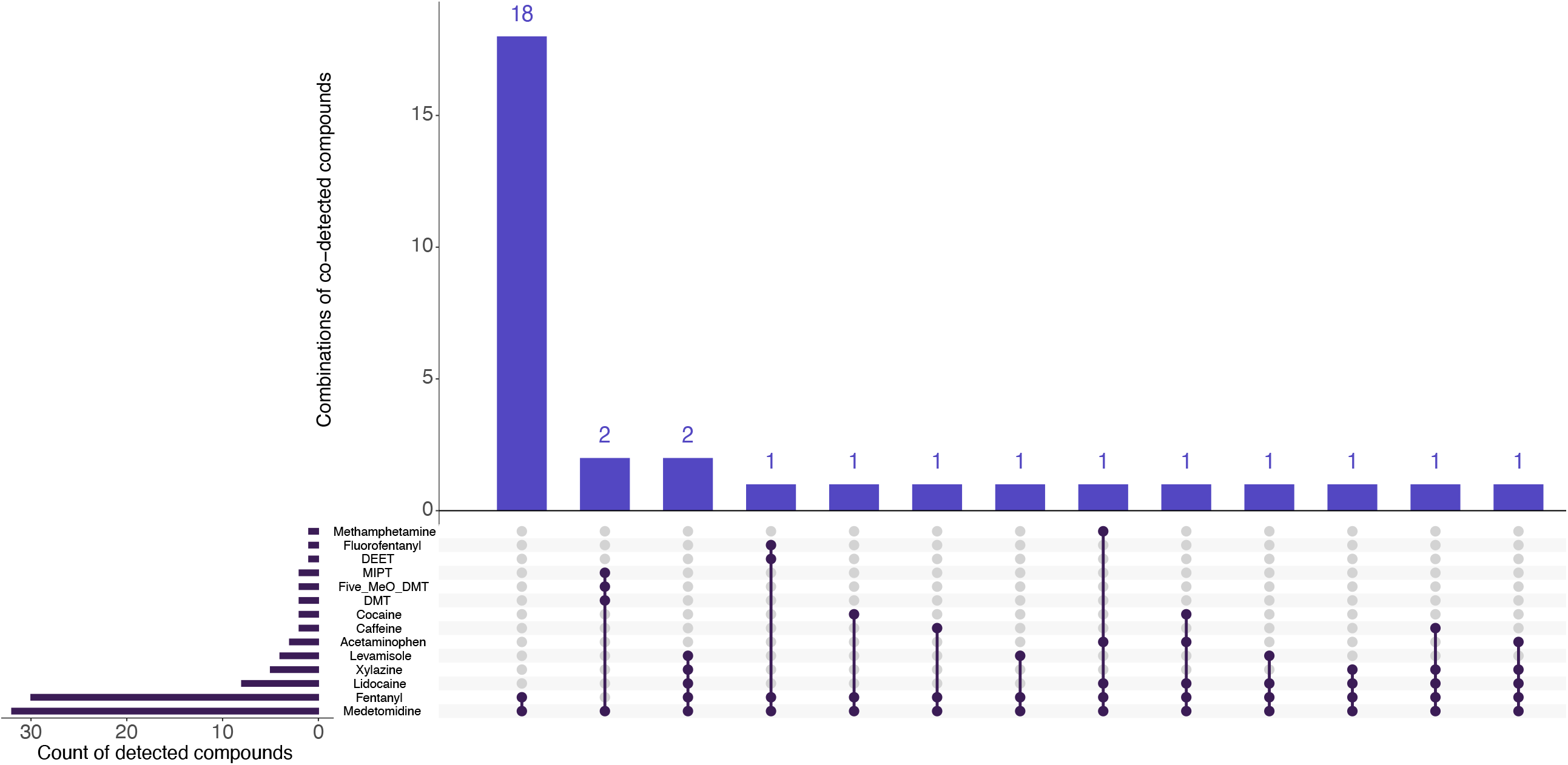
Drugs and adulterants co-present with sampled medetomidine in Southern California, 2025 - 2026. Note: N = 32 medetomidine-involved samples confirmed present on DART-MS. Data collected through a community-based drug checking program in Southern California, 2023 – 2026.

Medetomidine was detected in 30 (≈7 %) of 426 samples with fentanyl detected by DART-MS since it was first detected in May 2025. The quarterly prevalence of medetomidine co-present with fentanyl rose from 6 % (9 medetomidine-positive out of 149 fentanyl-positive) in Quarter 2, 2025 to 25 % (5 medetomidine-positive out of fentanyl-positive 20) by August of Quarter 3, 2026 (**Figure 1B**).

**Fig. 1B:**
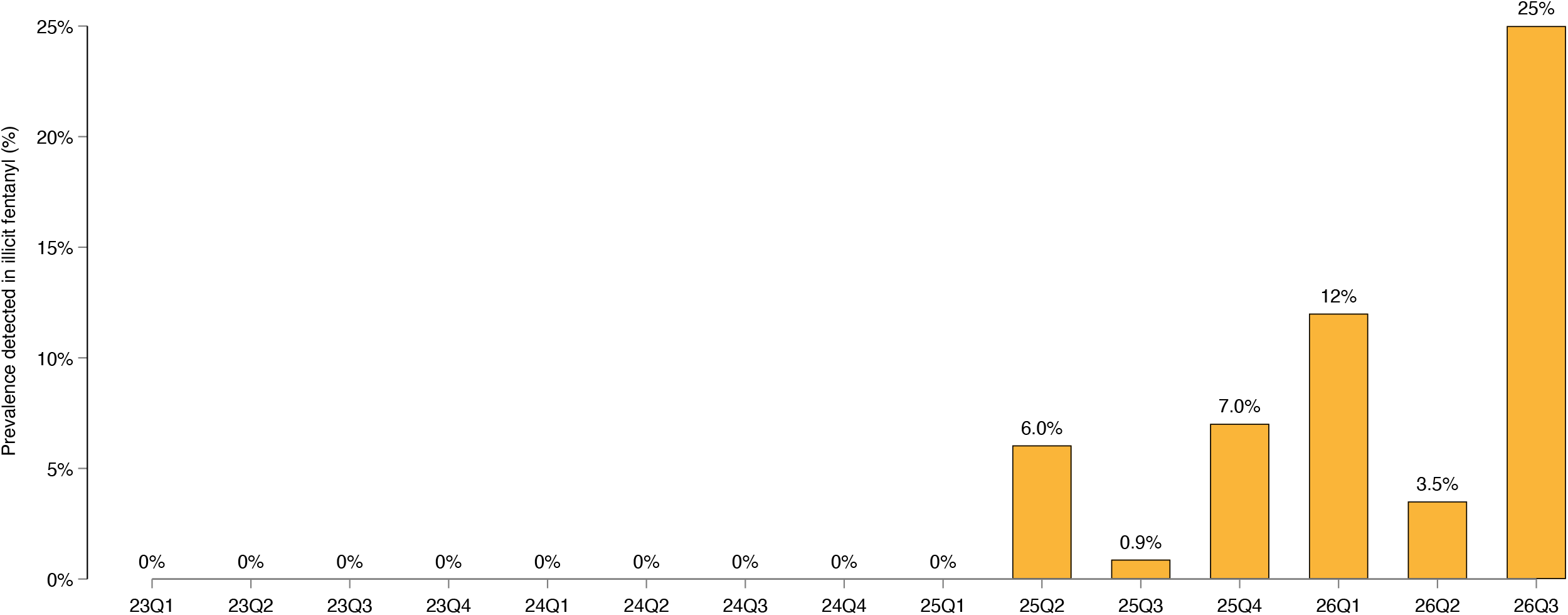
Prevalence of medetomidine detected in fentanyl in Southern California, by quarter, 2023 - 2026. Note: N = 30 fentanyl and medetomidine co-involved samples confirmed present on DART-MS; n = 2 samples of medetomidine co-detected with DMT excluded. Prevalence is calculated as the number of medetomidine-positive fentanyl samples divided by the total number of fentanyl-positive samples, per quarter. Data collected February 2023 – August 2026.

Surprisingly, two samples of expected dimethyltryptamine (DMT) obtained by a participant via the dark web were also confirmed to have medetomidine present, as well as another short-acting potent psychedelic, 5-methoxy-*N,N*-dimethyltryptamine (5-MeO-DMT), and the novel designer tryptamine methylisopropyltryptamine (MiPT) (**Figure 1A, Table 1**).

The mean concentration of medetomidine in the 22 quantitated samples was 0.4 % mass fraction, with a standard deviation of ±0.4 % mass fraction, and a range of below the limit of detection (<0.1 % mass fraction) to 1.4 % mass fraction. The presence of medetomidine in fentanyl was not associated with a statistically significant difference (p = .184) in the concentration of fentanyl according to pooled t-tests. Among the n = 13 samples where additional enantiomeric analysis was conducted, all thirteen samples were identified as containing a racemic mixture of medetomidine, i.e., approximately equal parts dexmedetomidine and levomedetomidine.

The location where samples were purchased or otherwise obtained had significant geographic dispersion across Southern California, including Los Angeles, Orange, and San Diego counties. Among n = 17 samples containing medetomidine where the participant answered they had already tried it, almost all (88 %, 15 out of 17) reported something strange about the subjective experience which could potentially be attributed to medetomidine. Some of these unexpected effects included unusually strong sedation and drowsiness that was distinct from an opioid “nod”, a strange chemical taste when smoked, and the perception that the sample was more potent. Some participants reported that due to its strength, samples that were found to contain medetomidine were sometimes dubbed and sold as “rhino dope” or “rhino tranq”. In a few instances, participants reported mild visual disturbances, comparing them with hallucinations normally associated with classic psychedelics like psilocybin and lysergic acid diethylamide (LSD).

## Discussion

Medetomidine represents a rapidly emerging adulterant in the Southern California illicit fentanyl supply. In the latest time period available (Quarter 3 2026), medetomidine was identified in about 25% of the fentanyl-involved samples at a drug checking program operating in Southern California, which is more than four times as prevalent than when medetomidine was first detected by the program in Quarter 2 2025. Fentanyl adulterated with medetomidine was obtained at multiple drug checking sites from various participants over the course of more than a year, indicating that medetomidine’s adulteration in fentanyl is more widespread than a single batch in a single neighborhood in a single season. Although Southern California’s lower prevalence of xylazine has been distinct from other areas in the United States (Friedman et al., 2025), it is difficult to say whether medetomidine adulteration will follow similar geospatial and temporal patterns.

Medetomidine’s growing notoriety as a sedative in illicitly manufactured fentanyl may have unexpected spillover into other drug markets. For example, the two instances of medetomidine that were detected alongside DMT mark the first instance in the scientific literature, to our knowledge, of DMT’s adulteration with the sedative. The use of medetomidine as an adulterant in these two cases may serve as an attempt to mimic the intense and short-acting immersive psychedelic experiences common with typical doses of DMT and 5-MeO-DMT while reducing costs associated with production.

The average medetomidine concentration in fentanyl was 0.4 % mass fraction. This level may be relevant for clinical outcomes, as medetomidine is estimated to be up to approximately 100 times more potent than xylazine in animal models (de Andrade Horn et al., 2024). Participants almost always noted significantly different and stronger subjective effects than with unadulterated fentanyl, further suggesting this small amount may be pharmacologically impactful and relevant for clinical outcomes. The finding that all thirteen samples that underwent enantiomeric analysis were racemic mixtures indicates that it was not diverted pharmaceutical dexmedetomidine. However, the diversion of veterinary medetomidine, which is also a racemic mixture, cannot be ruled out.

Future research should continue to understand the health effects of medetomidine in the context of illicitly manufactured fentanyl, particularly the role of withdrawal symptoms, the potentiation of opioid effects, particularly those on the central nervous system and breathing, and any impact on clinical management of overdose. Toxicologic panels where fentanyl is identified or suspected, including hospital overdose response and post-mortem toxicology, should consider including medetomidine as a part of routine surveillance. Public health and harm reduction organizations could consider expanding access to technologies that identify medetomidine, such as low-cost medetomidine immunoassay test strips. Identifying medetomidine as an adulterant can help people who use fentanyl mitigate the risk of negative or unexpected effects (e.g., starting low and going slow, using in a safe environment around trusted others, having naloxone available) or to avoid medetomidine-adulterated fentanyl altogether.

### Limitations

There are several limitations that should be considered. First, our sample may have significant convenience bias and is therefore may not be representative of the broader Southern California drug market. For example, a participant may have decided to have their drugs checked only after there was an unusual experience. While we aimed to collect samples from geographically and culturally distinct locations and spaces across the county, the samples brought to drug checking may be systematically different than those that were not. Drug checking services were held primarily in or near syringe service programs and harm reduction drop-in centers, which may have also biased the results. Laboratory methods may be subject to various errors.

## Conclusion

Medetomidine continues to be an emerging sedative adulterant, newly identified in Southern California illicit fentanyl markets in clinically relevant amounts. The introduction of medetomidine adulteration in these markets may change use patterns and preferences for α2 sedative adulteration in illicitly manufactured fentanyl. Future research could explore how medetomidine affects overdose, withdrawal, and short and long-term health more generally. The ability for surveillance and monitoring systems to identify and track the presence of medetomidine in health systems and death records is paramount for public health.

## Data Availability

Individual line-level data produced in the present study are unavailable. Aggregated data is available online at drugcheckinglosangeles.com/viz

## Funding Acknowledgements

This work was supported by the Centers for Disease Control and Prevention as part of Overdose Data to Action: LOCAL (NH28CE003543-01-00), National Institutes of Health (R01 DA057630), and an equipment grant from the James B. Pendleton Charitable Trust to the UCLA AIDS Institute and UCLA Center for AIDS Research. AJK was supported by the NIH/National Center for Advancing Translational Science (NCATS) UCLA CTSI (grant TL1TR001883). A portion of this work is funded by the Division of Overdose Prevention, National Center for Injury Prevention and Control (NCIPC), Centers for Disease Control and Prevention. The funders had no role in designing the study, analyzing the data, or the decision to submit for publication.

## Disclaimer

Certain commercial products, instruments, software, or materials are identified to adequately specify the procedure; this does not imply endorsement or recommendation by NIST, nor does it imply that such products are necessarily the best available for the purpose. These opinions, recommendations, findings, and conclusions do not necessarily reflect the views or policies of NIST or the United States Government.

## Additional Acknowledgments

We would also like to acknowledge our community partners and volunteers who assisted in field services and sample collection. Lastly, we would like to extend our gratitude to the anonymous participants of Drug Checking Los Angeles who made this work possible through their donation of samples for confirmatory testing.

## Notes

### Competing Interest Statement

The authors have declared no competing interest.

### Author Declarations

The University of California Los Angeles Institutional Review Board of the University of California Los Angeles gave ethical approval for this work under protocola (IRB-22-0760 and IRB-22-1273) and also determined some components of this work constituted public health surveillance. The National Institute of Standards and Technology Research Protections Office of the National Institute of Standards and Technology gave ethical approval for this work (MML-2021-0328).

