## Supplemental Table A for "Characterizing the Emergence of Medetomidine in Southern California Fentanyl Markets"

| **Supplemental table A: LC/MS Quantitation Panel** | |
| --- | --- |
| **Class** | **Quantitative Analyte** |
| **α_2_- agonists** | Medetomidine |
|  | Xylazine |
| **Fentanyl, fentalogs, and fentanyl precursors** | Fentanyl |
|  | Carfentanil† |
|  | Fluorofentanyl |
|  | 4-ANPP |
|  | Ethyl 4-ANPP |
|  | Phenethyl 4-ANPP |
| **Anesthetics** | Cocaine |
|  | Lidocaine* |
|  | Procaine† |
|  | Tetracaine |
| **Heroin and related compounds** | Heroin |
|  | 6-MAM† |
|  | Noscapine† |
|  | Papaverine† |
| **Amphetamines** | Methamphetamine |
| **Other drugs** | Acetaminophen |
|  | Caffeine† |
|  | Ketamine |
|  | Levamisole† |
|  | Quetiapine (Seroquel) |
| **Cutting agents & fillers** | BTMPS* |
| *Added to the quantitation panel in July 2024  †Added to the quantitation panel in January 2025 | |
